# Multimodal Deep Learning for Longitudinal Prediction of Glaucoma Progression Using Sequential RNFL, Visual Field, and Clinical Data

**DOI:** 10.1101/2025.10.31.25339266

**Authors:** Mousa Moradi, Jerry Cao-Xue, Mohammad Eslami, Mengyu Wang, Tobias Elze, Nazlee Zebardast

## Abstract

Forecasting glaucoma progression remains a major challenge in preventing irreversible vision loss. We developed and validated a multimodal, longitudinal deep learning framework to predict future progression using a large retrospective cohort of 10,864 patients from Mass Eye and Ear. The model integrates sequential structural (OCT RNFL scans), functional (visual-field maps), and clinical data from a two-year observation window to forecast progression over the subsequent two-to four-year horizon. Four backbone architectures (ConvNeXt-V2, ViT, MobileNet-V2, EfficientNet-B0) were coupled with a bidirectional LSTM to capture temporal dynamics. The ConvNeXt-V2-based model achieved 0.97 AUC and 0.94–0.96 accuracy, outperforming other backbones with robust performance across sex and race subgroups and only modest attenuation in those > 70 years. Saliency maps localized to clinically relevant arcuate bundles, supporting biological plausibility. By effectively fusing multimodal data over time, this framework enables accurate, interpretable, and equitable long-horizon risk stratification, advancing personalized glaucoma management.

## I. INTRODUCTION

Glaucoma is a leading cause of irreversible blindness worldwide, affecting more than 70 million people, with numbers projected to exceed 110 million by 2040 as populations age [1]. The disease is marked by progressive loss of retinal ganglion cells, which manifests structurally as thinning of the retinal nerve fiber layer (RNFL) and functionally as visual field (VF) deterioration measured by standard automated perimetry (SAP) [2]. Because neuronal damage is permanent once established, the timely identification of patients at risk of rapid progression is a central challenge in glaucoma management.

Current clinical practice relies on longitudinal monitoring of RNFL thickness using optical coherence tomography (OCT) and VF mean deviation indices. However, regression-based slope analyses of these parameters are noisy, require years of follow-up to reach statistical significance, and are sensitive to measurement variability [3, 4]. Moreover, real-world data are characterized by irregular visit intervals, missed follow-ups, and patient attrition, which further complicates robust estimation of progression trajectories [5, 6]. As a result, patients may only be recognized as progressors after substantial and irreversible vision loss has occurred.

Artificial intelligence (AI) has emerged as a promising tool to address these challenges. Deep learning models have achieved expert-level performance in ophthalmology, particularly in disease detection from fundus photography and cross-sectional OCT scans [7, 8]. Temporal models, including recurrent neural networks and transformers, have shown potential to capture longitudinal dependencies in clinical and imaging data [9, 10]. Multimodal strategies that integrate complementary structural and functional signals, such as RNFL thinning and VF deterioration, have been proposed to improve prediction accuracy [11].

Despite these advances, most published work in glaucoma AI has focused on cross-sectional diagnosis or narrowly defined prediction tasks, rather than robust long-horizon progression forecasting. For example, Yousefi et al. (2014) combined RNFL thickness from OCT and SAP VF features as tabular data using classical classifiers (Random Forest, Bayes Net), achieving an AUC of 88% in predicting progression within two years [12]. Dixit et al. (2021) applied convolutional LSTMs to longitudinal VF sequences of pointwise sensitivity values from over 11,000 eyes, reporting accuracies of 91–93% across four visits, but their model was restricted to functional data alone [13]. Tarcoveanu et al. (2022) evaluated multiple classifiers on small datasets (~150 eyes) using clinical variables such as intraocular pressure, VF indices (mean deviation (MD), pattern standard deviation), and RNFL values, reporting accuracies around 92% but with limited generalizability [14]. More recently, Afolabi et al. (2025) introduced FairDist, an equity-aware EfficientNet model trained exclusively on OCT B-scans (100,000 images from 500 subjects) without incorporating VF data, yet performance remained modest (AUC = 74%) despite the use of fairness-aware learning [15]. Addressing these limitations requires frameworks that integrate structural, functional, and clinical-demographic information over time, enabling robust long-horizon prediction.

To address these gaps, we assembled the largest multimodal glaucoma dataset to date (>10,000 patients; 149,000 paired RNFL OCT and VF images with clinical covariates) and developed a deep learning framework that integrates OCT, VF, and clinical features for long-horizon progression prediction. We systematically evaluated four state-of-the-art backbones (ConvNeXt [16], Vision Transformer (ViT) [17], MobileNet [18], and EfficientNet [19]) within a Bi-LSTM temporal fusion architecture. The proposed model achieved state-of-the-art performance (94% accuracy, 95% AUC) while maintaining fairness and interpretability across demographic subgroups. Overall, our contributions are as follows:

1. We introduce a large-scale, longitudinal multimodal dataset combining OCT, VF, and clinical data for glaucoma progression analysis.
2. We propose a Bi-LSTM-based deep learning framework for multimodal temporal fusion, demonstrating superior performance and demographic fairness.
3. We provide interpretability and uncertainty analyses showing that model attention aligns with clinically relevant structural-functional regions, supporting transparent clinical adoption.

This study presents a deep learning model for accurate and equitable glaucoma progression prediction. **Section I** introduces the clinical context and prior methods. **Section II** describes dataset construction, preprocessing, and model architecture. **Section III** outlines implementation details, comparative and ablation experiments. **Section IV** discusses findings and implications, and **Section V** concludes with future directions for multimodal longitudinal AI in ophthalmology.

## II. Materials and Methods

### A. Study Population, Cohort Construction, and Label Definition

The Institutional Review Board at Mass General Brigham (MGB) approved this study, which adhered to the ethical guidelines outlined in the Declaration of Helsinki for research involving human participants. We conducted a retrospective longitudinal cohort study of glaucoma patients seen at Massachusetts Eye and Ear (MEE) between 2010 and 2023. Given the retrospective design, the requirement for informed consent was waived. This yielded in 26,562 patients with 217,041 VFs and 46,963 patients with 320,221 RNFL OCT scans. Glaucoma patients were identified using the PyGlaucoMetrics [20] package, and eyes were classified as glaucomatous if at least two reliable 24-2 VF tests were available on different dates. Patients were included if they had at least two reliable VF tests and corresponding RNFL OCT scans within the first two years after baseline, with follow-up extending at least three years. VF and OCT records were merged by patient ID, eye laterality, and exam date, and visits were matched if VF and RNFL exams occurred within a ±6-month window to ensure temporal alignment. Reliable exams were defined as 24-2 VF tests with a false-positive rate < 33% and RNFL OCT scans with signal strength ≥ 7. RNFL thickness was measured from Circumpapillary retinal nerve fiber layer (cpRNFL) around the optic nerve head using the Zeiss Cirrus OCT (Carl Zeiss Meditec, Inc., Dublin, CA, USA) device. Demographic and clinical covariates, including age, sex, race, mean deviation (MD), and average RNFL thickness, were extracted from the metadata and incorporated into the dataset. The final matched cohort consisted of 10,864 patients with 149,612 longitudinal VF-RNFL pairs and corresponding clinical data. An observation window of two years after baseline was used to collect input sequences, with up to three timepoints selected using nearest-neighbor matching at approximately 0, 1, and 2 years. The prediction horizon was defined as 2-4 years after baseline to account for irregular follow-up schedules. This interval was chosen because glaucoma progression is typically detectable within 2-5 years (Fig. 1) of follow-up under routine clinical care [4]. Eyes were labeled as “progressors” using the vfprogression [21] package if VF progression analysis indicated statistically significant worsening within this prediction window, and as “stable” otherwise. We selected Visual Field Index (VFI) over MD slope for progression definition because VFI preferentially weights central visual field points and minimizes the influence of diffuse depression, making it more robust to media opacity changes [22]. Notably, studies have demonstrated that while MD and pattern standard deviation (PSD) show significant changes following cataract extraction, VFI remains relatively stable, indicating superior resistance to lenticular-induced artifacts in longitudinal progression analysis [23].

**Fig. 1.**
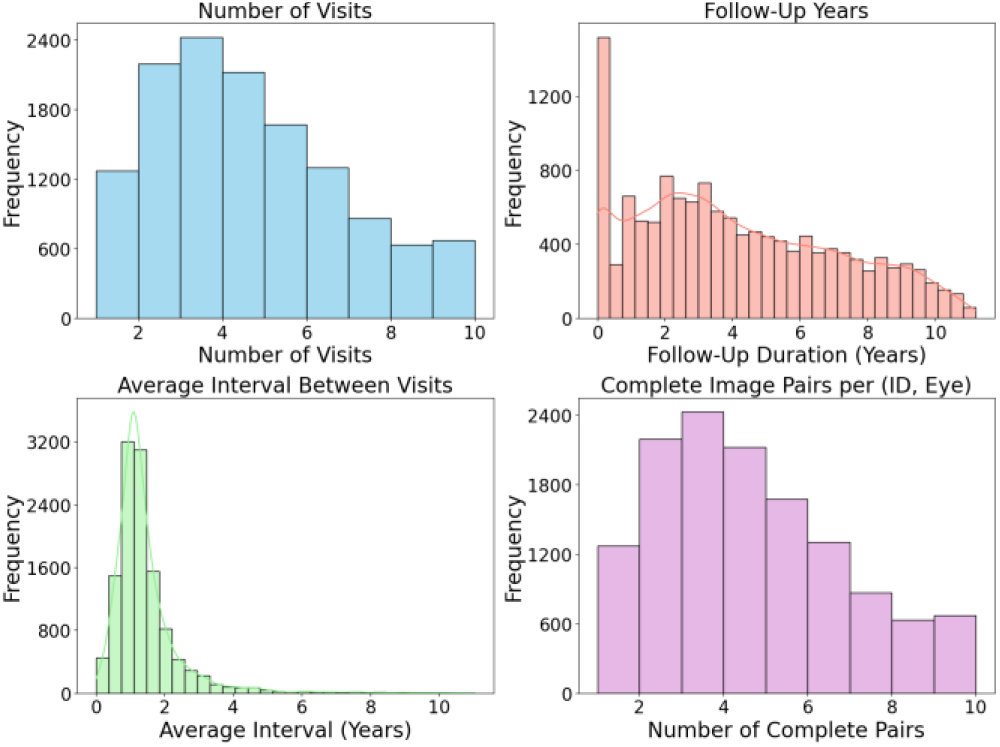
Histogram of time-series data evaluated for this study.

### B. Visual Field Image Construction

Standard automated perimetry tests were obtained using the Humphrey Field Analyzer (HFA II; Carl Zeiss Meditech, Dublin, CA, USA). Pointwise sensitivities were extracted from HFA 24-2 VF tests, which are widely regarded as the clinical standard for glaucoma diagnosis and follow-up [24]. From these tests, TD values were computed for all 52 test locations, excluding the two blind spots, using the PyGlaucoMetrics [20] package 16. To enable image-based deep learning, we spatially mapped the 52 TD values onto an 8 × 9 matrix corresponding to the HFA 24-2 test grid. Blind spot positions were masked and set to NaN values. The resulting VF TD maps were treated as grayscale images, with pixel intensities representing deviation values in decibels. This approach preserved the spatial topology of VF loss while allowing integration with RNFL OCT images in the multimodal framework.

### C. Data preprocessing

All VF and RNFL images were resized to 224×224 pixels and normalized to ImageNet statistics (mean [0.485, 0.456, 0.406], standard deviation [0.229, 0.224, 0.225]) to ensure compatibility with pretrained convolutional backbones. Images were stored in RGB format, and intensity scaling was applied to reduce variability across acquisition sessions. Tabular features were preprocessed separately: categorical variables such as race were one-hot encoded, and continuous features including age, MD, and RNFL thickness were z-score normalized using a fold-specific scaler to avoid information leakage. Missing visits within the observation window were handled using nearest-neighbor matching in time, such that each expected timepoint (~0, ~1, ~2 years) was paired with the closest available examination within a ±6-month tolerance.

#### Algorithm 1 contructing VF TD Images from HFA 24-2 Tests

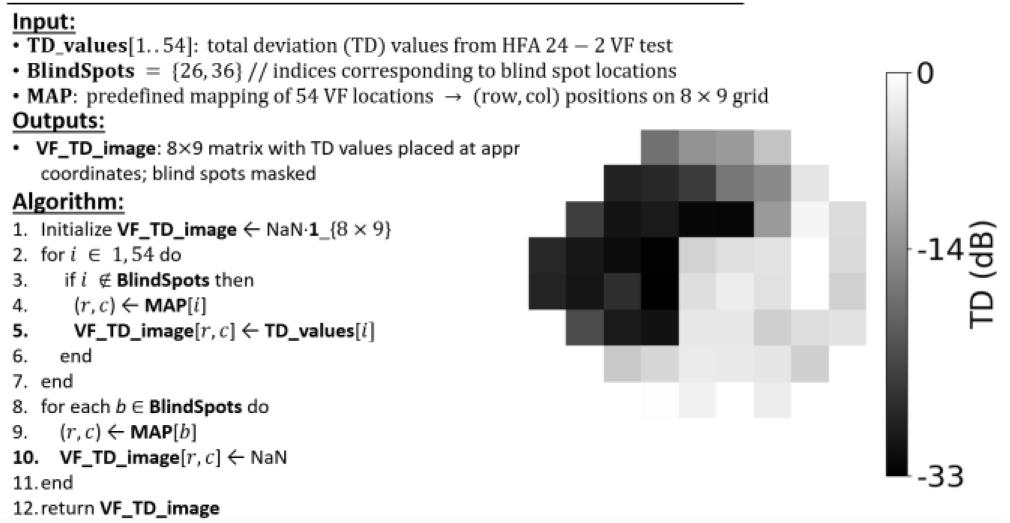

### D. Model Architecture

We developed a multimodal deep learning model that jointly processes VF images, RNFL images, and tabular clinical features. For rigorous analysis, we trained four architectures (ConvNeXt-V2 [16], Vision Transformer [17] (ViT-base), EfficientNet-B0 [19], and MobileNet-V2 [18]) using the same input data and hyperparameters. For each backbone, RNFL and VF images were encoded with shared weights, while clinical features were processed by a two-layer fully connected encoder (64 to 128 dimensions). At each timepoint, encoded RNFL, VF, and tabular features were concatenated into a fused feature vector. Temporal dynamics across visits were modeled with a two-layer bidirectional long short-term memory (Bi-LSTM) network with 256 hidden units and dropout of 0.3. The final hidden state was passed to a classifier composed of a dense layer (128 units, ReLU activation) and a sigmoid output for binary progression prediction. Hyperparameters were tuned using the Optuna [25] with 50 trials, with the search space including learning rate (2 × 10^−5^), batch size (16), weight decay (1 × 10^−2^), warm-up epochs (10), training epochs (200), and early stopping patience (11). Figure 1 shows the general block diagram for the proposed longitudinal study.

### E. Statistical Analysis and Performance Metrics

Model performance was primarily assessed using area under the receiver operating characteristic curve (AUC). Secondary metrics included F1 score, accuracy, and precision–recall AUC. Confusion matrices were aggregated across folds to evaluate classification balance. Results were reported as mean ± standard deviation across five folds. In addition, gradient-weighted class activation maps (Grad-CAMs [26]) were computed to visualize salient image regions contributing to predictions. Prediction uncertainty for each sample was determined using Tsallis entropy [27], computed as

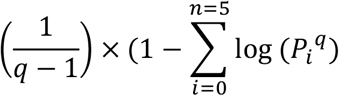

where *P*_*i*_ is the predicted probability of class *i*. Given the class imbalance, we set the entropic parameter to *q* = 0.25 which provides greater sensitivity to progression predictions [28].

All tests were two-sided with significance set at *P* value< 0.05. Model performance was compared across metrics, data fractions, and demographic subgroups. Pairwise differences (accuracy, F1, AUROC, AUCPR, PV score) were assessed using paired t-tests across five cross-validation folds. Subgroup analyses (race, gender, age) used ANOVA with post-hoc t-tests. P-values were adjusted for multiple comparisons using Bonferroni correction where appropriate. Analyses were performed in Python 3.10 using SciPy (v1.13) and statsmodels (v0.14).

## III. Results

### A. Patient Demographics and Clinical Characteristics

Table I illustrates baseline characteristics between progressors (N=7,779 eyes) and non-progressors (N=31,887 eyes). Progressors were significantly older (70.5±12.8 vs. 65.6±14.7 years), had similar gender distribution (51.1% vs. 58% female) but different racial composition, with White patients representing the majority in both groups (69.6% vs. 63.5%, *P* value < 0.001). Progressors demonstrated worse baseline disease with lower MD (−8.11±6.24 vs. −5.9±5.25 dB), thinner RNFL (59.4±9.5 vs. 77.9±11.3 μm), and faster deterioration rates for both MD slope (−0.013 vs. −0.005 dB/year) and RNFL slope (−0.530 vs. −0.264 μm/year) (all *P* values < 0.001).

### B. Performance Evaluation and Ablation Experiments

Across five-fold cross-validation, our multimodal framework demonstrated robust predictive performance, with ConvNeXt consistently outperforming other backbones across all demographic subgroups. Overall, ConvNeXt achieved the highest accuracy (0.947) and F1-score (0.887), followed by ViT (accuracy 0.938, F1 0.838), MobileNet (accuracy 0.723, F1 0.752), and EfficientNet (accuracy 0.329, F1 0.325), with all pairwise comparisons statistically significant (*P* value < 0.001).

Race-stratified analyses (Fig. 2a) revealed consistent performance across racial groups. ConvNeXt maintained superior accuracy (0.94-0.95) and F1-scores (0.88-0.89) across Asian, Black, and White patients, with no significant performance differences between groups (P > 0.1). ConvNeXt exhibited the lowest prediction uncertainty (0.195), significantly lower than ViT (0.261), MobileNet (0.270), and EfficientNet (0.908) (all *P* value < 0.01).

**Fig. 2.**
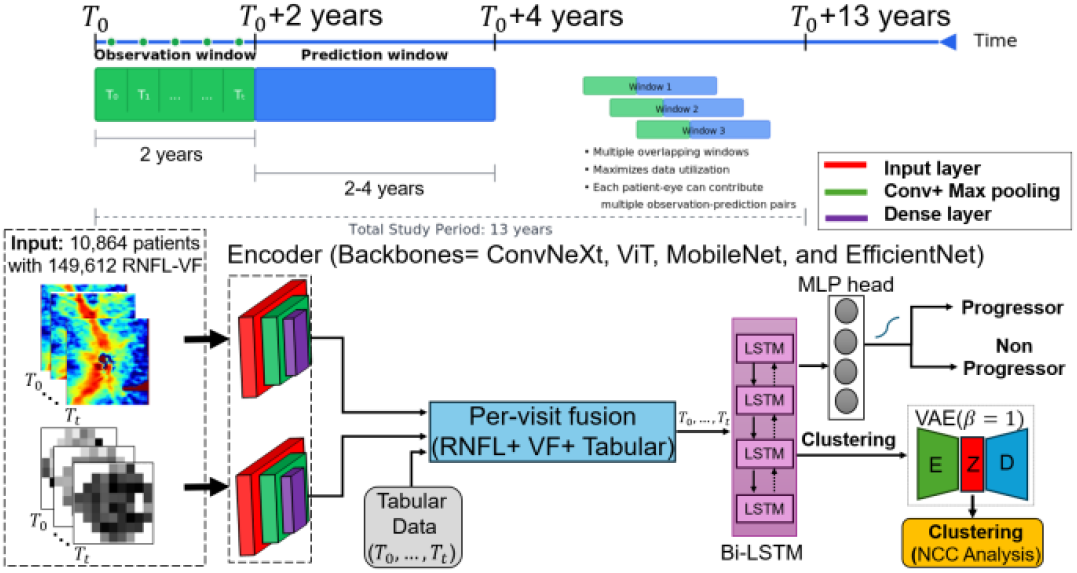
Study design and multimodal deep learning framework for glaucoma progression prediction. Each training instance consisted of a 2-year observation window (T_0_…T_t_) followed by a 2-4 year prediction horizon, with overlapping sliding windows to maximize data utilization. At each visit, RNFL and VF images were encoded by convolutional neural network backbones, and structured tabular features (age, sex, race, global RNFL thickness, VF mean deviation) were projected through a multilayer perceptron. The resulting embeddings were concatenated into a per-visit fused representation and passed through a two-layer bidirectional LSTM to capture temporal dynamics. The final hidden state was processed by a dense head with sigmoid output to classify eyes as progressors or non-progressors.

Sex-stratified analyses (Fig. 2b) demonstrated balanced performance between sexes. ConvNeXt and ViT achieved comparable high accuracy (>0.93) in both female and male subgroups, with ConvNeXt maintaining F1-scores 0.87-0.89 and ViT 0.83-0.85. No significant sex-related performance differences were observed (*P* value> 0.1). Prediction uncertainty remained lowest for ConvNeXt (0.195-0.202) across both groups.

Age-stratified analyses (Fig. 2c) revealed performance degradation with advancing age, particularly for MobileNet. In patients >70 years, MobileNet performance dropped substantially (accuracy 0.468, F1 0.473), while ConvNeXt maintained robust performance (accuracy 0.943, F1 0.868) and ViT showed modest decline (accuracy 0.930, F1 0.823). ConvNeXt demonstrated the most stable uncertainty across age groups (0.195-0.200), indicating consistent reliability regardless of patient age.

**TABLE 1.** PATIENT DEMOGRAPHICS AND CLINICAL CHARACTERISTICS. CONTINUOUS VARIABLES ARE PRESENTED AS MEAN ± SD. CATEGORICAL VARIABLES ARE PRESENTED AS N (%). INDEPENDENT SAMPLES T-TESTS FOR CONTINUOUS VARIABLES AND CHI-SQUARE TESTS FOR CATEGORICAL VARIABLES. STATISTICAL SIGNIFICANCE WAS SET AT P < 0.05. NS=NOT SIGNIFICANT.

| Characteristic | Progressors<br>(7779 eyes) | Non-progressors<br>(31887 eyes) | P-value |
| --- | --- | --- | --- |
| Age at baseline, years | 70.5±12.8 | 65.6±14.7 | <0.001 |
| Gender, n(%) |  |  |  |
| Female | 3972(51.1) | 18497(58) |  |
| Male | 3807(48.9) | 13390(42) |  |
| Race, n(%) |  |  | <0.001 |
| Asian | 593(7.6) | 2398(7.5) |  |
| Black/African American | 1148(14.8) | 5348(16.8) |  |
| White | 5414(69.6) | 20236(63.5) |  |
| Other | 624(8) | 3905(12.2) |  |
| Follow-up time, years | 2.1± 1 | 2.1± 1 | ns |
| Mean TD at baseline, dB | -7.82±5.98 | -5.78±5.08 | <0.001 |
| Mean MD at baseline, dB | -8.11±6.24 | -5.9±5.25 | <0.001 |
| MD slope at baseline, dB/year | -0.013 | -0.005 | <0.001 |
| Mean RNFL at baseline, μm | 59.4±9.5 | 77.9±11.3 | <0.001 |
| RNFL slope at baseline, μm/year | -0.530 | -0.264 | <0.001 |

### C. Feature Importance Maps and Contribution Analysis

Gradient-weighted class activation maps averaged across five-fold cross-validation revealed distinct patterns of feature utilization across architectures for both RNFL OCT and VF TD images (Fig. 3). ConvNeXt consistently highlighted clinically relevant regions, including the superior and inferior arcuate bundles in RNFL OCT scans and corresponding arcuate and paracentral clusters in VF images, reflecting the known anatomical correspondence between structural damage and functional loss in glaucoma. ViT also captured meaningful regions, but with more diffuse and less localized importance, suggesting reduced precision in aligning structural and functional cues. MobileNet and EfficientNet demonstrated noisy and spatially inconsistent activation patterns, often emphasizing background or non-informative regions in both OCT and VF maps, consistent with their lower predictive performance.

**Fig. 3.**
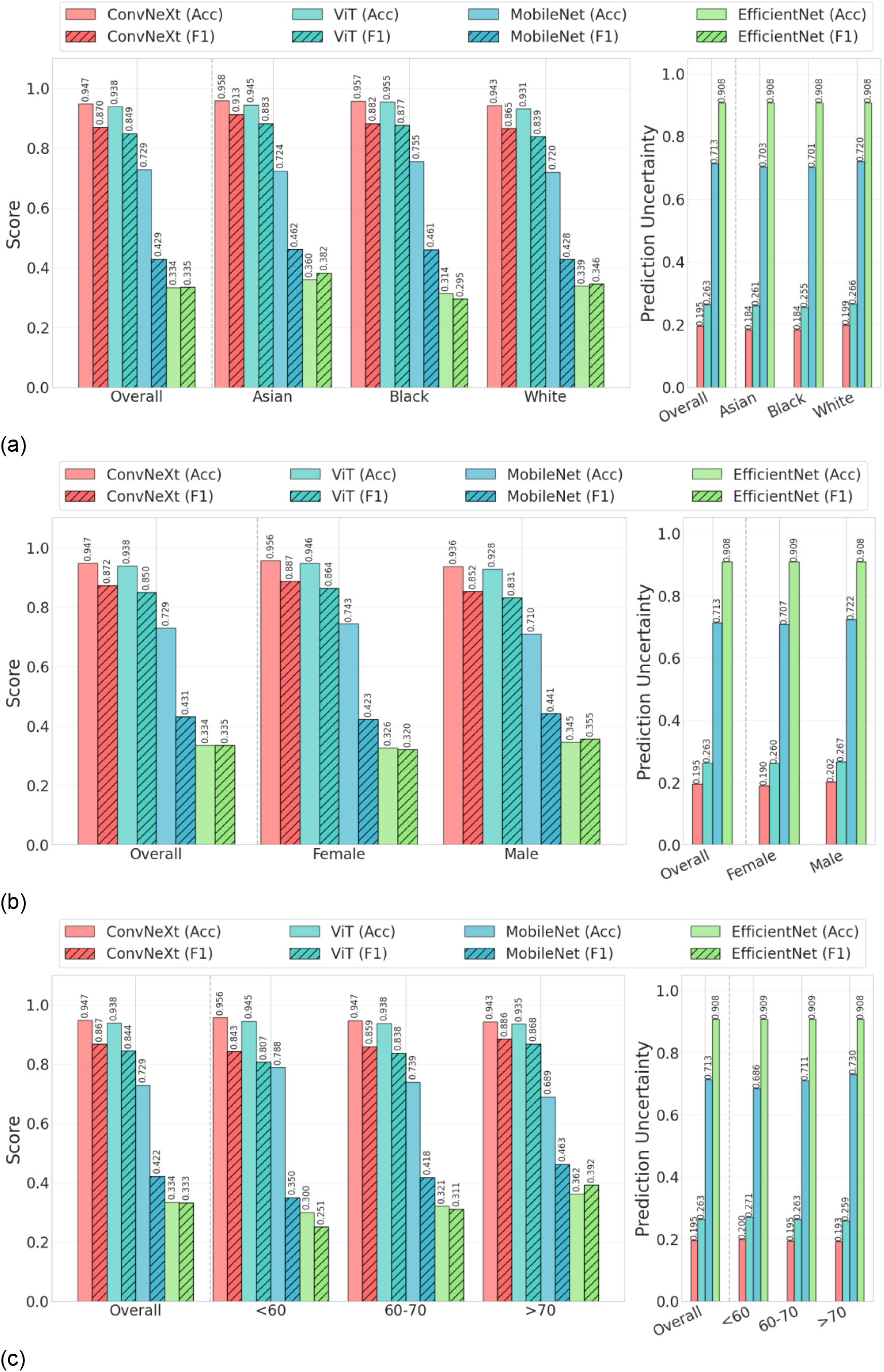
Performance metrics and uncertainty analysis across demographic subgroups. Comparison of four backbone architectures (ConvNeXt, Vision Transformer, MobileNet, EfficientNet) in five-fold cross-validation. **a** Race-stratified analyses, **b** Sex-stratified results, **c** Age-stratified analyses. EfficientNet consistently underperformed with high uncertainty values (~0.909, *P* value < 0.001).

**Fig. 4.**
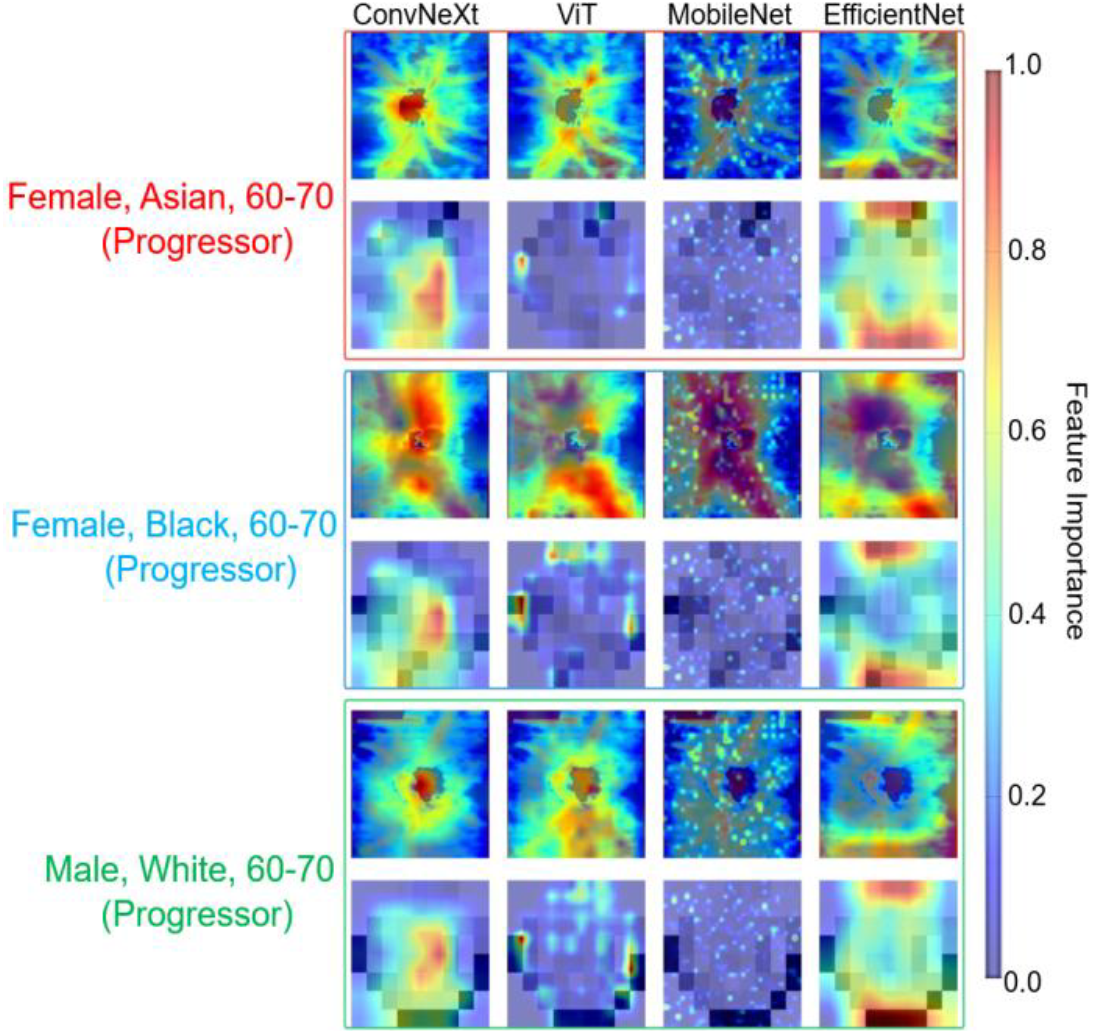
Gradient-weighted activation maps (Grad-CAMs) for RNFL OCT and VF TD images across backbones and subgroups. Averaged over five folds, ConvNeXt consistently focused on clinically relevant arcuate regions in both modalities, while ViT produced diffuse and MobileNet/EfficientNet noisy activations. Patterns remained stable across demographic subgroups, supporting the robustness and fairness of the ConvNeXt framework.

Importantly, these findings were consistent across demographic subgroups (Female Asian, Female Black, Male White, ages 60-70), indicating that ConvNeXt’s strong performance was not driven by spurious features or subgroup-specific biases. Instead, the model leveraged pathophysiologically plausible features across populations, supporting both the accuracy and fairness of our multimodal framework.

As summarized in Table 2, our multimodal model clearly outperformed prior approaches for glaucoma progression prediction. Earlier machine learning methods, such as Yousefi et al. [12], achieved an AUC of 88% on a small cohort of 180 eyes using RNFL and VF data, while Dixit et al. [13] reported accuracies of 91-93% (AUC= 89-93%) using longitudinal VF sequences and clinical data. Tarcoveanu et al. [14] tested multiple classifiers on very limited datasets (~150 eyes) and obtained 92% accuracy, though without AUC reporting. More recently, in 2025, Afolabi et al. [15] introduced FairDist with an EfficientNet backbone, but performance remained modest (AUC= 74%) despite using 500 subjects with 100,000 OCT B-scans. By contrast, our model, trained on the largest dataset to date (10,864 subjects; 149,612 multimodal RNFL–VF images), achieved 94% accuracy and 95% AUC over a 4-year prediction window, representing the best balance of scale, modality integration, and predictive performance among existing studies.

**TABLE 2.** BENCHMARKING OUR MODEL AGAINST STATE-OF-THE-ART GLAUCOMA PROGRESSION PREDICTION METHODS. THE TABLE SUMMARIZES DATASET SIZE, INPUT MODALITIES, PREDICTION WINDOWS, AND PERFORMANCE METRICS (ACCURACY AND AUC. IF VALUES ARE NOT REPORTED, THEY SHOW AS N/A= NOT APPLICABLE. BOLD VALUES SHOW THE BEST SCORES

| Author, year | Dataset | Model | Prediction window (years) | Accuracy (%) | AUC (%) |
| --- | --- | --- | --- | --- | --- |
| Yousefi et al., 2014 [12] | 180 eyes (107 progressing, 73 stable) , longitudinal OCT RNFL + VF data | Random Forest, Bayes Net, CART, etc. | 2 | N/A | 88 |
| Dixit et al., 2021 [13] | 11,242 eyes (longitudinal VF + clinical) | Convolutional LSTM | 4 | 91-93 | 89-93 |
| Tarcoveanu et al., 2022 [14] | 50 subjects (100 eyes, 3 visits) + 101 eyes (with diabetes) | Random Forest, MLP, kNN, SVM, etc. | 3 | 92 | N/A |
| Afolabi et al., 2025 [15] | 500 subjects with 100,000 OCT B-scans | FairDist with EfficientNet backbone | 6 | N/A | 74 |
| **Our model, 2025** | 10,864 subjects (39,666 eyes) with 149,612 RNFL-VF images | Feature fusion with ConvNext, ViT, MobileNet, and EfficientNet backbones | 4 | **94** | **95** |

Ablation studies revealed the contribution of each modality to model performance (Fig. 5). The full multimodal ConvNeXt model achieved exceptional performance (AUC 0.975±0.015), comparable to variants excluding tabular data (AUC 0.976±0.014) or using only RNFL (AUC 0.975±0.014) or VF (AUC 0.974±0.012) imaging. However, removing RNFL data caused dramatic performance degradation (No RNFL ConvNeXt, AUC 0.569±0.015), while removing VF data maintained high performance (No VF ConvNeXt, AUC 0.975±0.015), suggesting RNFL provides the critical structural information. The tabular-only model performed poorly (AUC 0.526±0.064), confirming imaging superiority over clinical variables alone. ViT showed similar patterns with strong full model performance (AUC 0.965±0.012) but substantial degradation without RNFL (AUC 0.752±0.106). MobileNet and EfficientNet consistently underperformed, with EfficientNet showing the poorest discrimination (full model AUC 0.607±0.016).

**Fig. 5.**
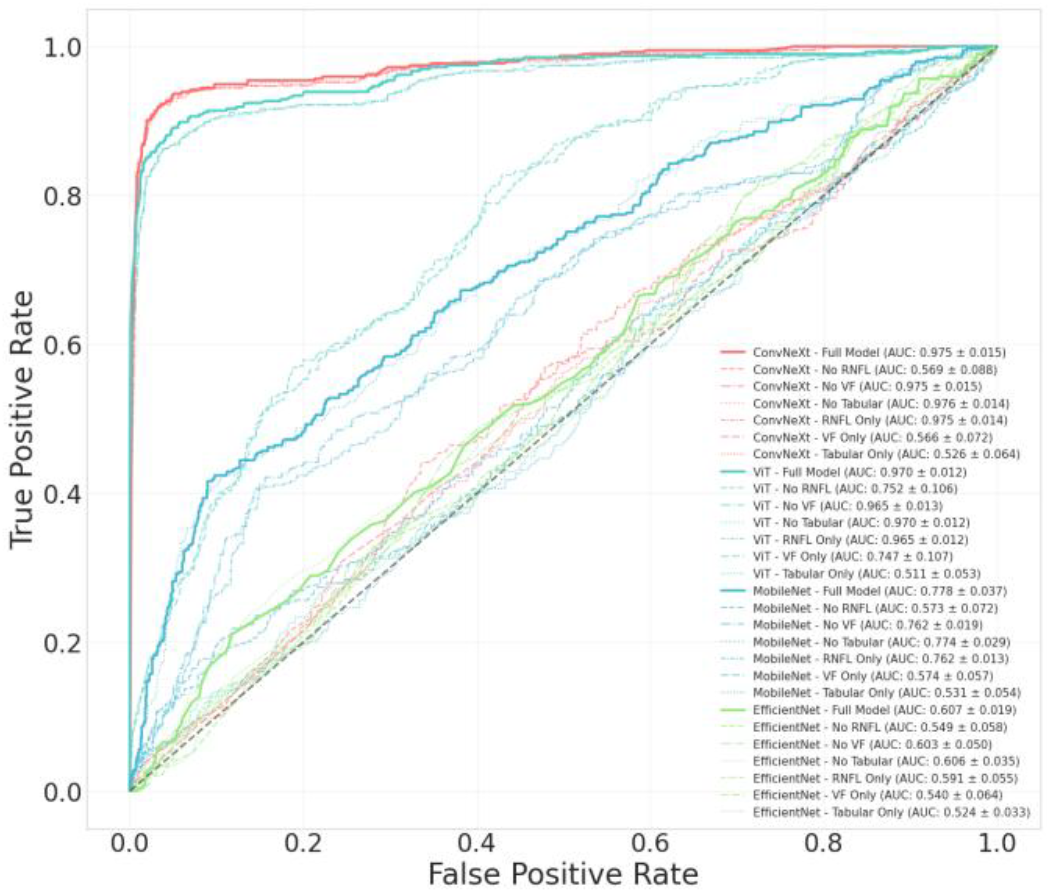
ROC curves demonstrating the contribution of multimodal features to glaucoma progression prediction. The full multimodal ConvNeXt model (AUC = 0.975±0.015) achieves near-perfect discrimination.

## IV. DISCUSSION

Our study advances glaucoma progression modeling beyond prior cross-sectional or single-modality efforts. Earlier multimodal systems achieved only moderate discrimination on small datasets, while VF-only sequence models ignored OCT structure and OCT-only networks lacked generalizability. We address these gaps through scale, fusion, and temporal modeling, leveraging an order-of-magnitude larger paired dataset (10,864 subjects; 149,612 RNFL-VF images), integrated multimodal inputs, and a Bi-LSTM encoder for longitudinal dynamics. Benchmarking against prior work (Table II) shows that earlier approaches reported AUC ≈ 88-93% on limited cohorts, whereas our multimodal, sequence-aware framework achieved AUC 95% and accuracy 94% over a 4-year horizon, underscoring the benefits of data scale and explicit structure–function fusion for long-term forecasting.

Clinically, early identification of likely progressors could enable risk-stratified follow-up, targeted testing, and proactive therapy. Translation should include calibration and decision-curve analyses to align model thresholds with local prevalence and harm-benefit trade-offs.

Grad-CAM maps consistently emphasized RNFL arcuate bundles and VF arcuate/paracentral zones, lending biological plausibility and explaining ConvNeXt’s advantage over ViT, MobileNet, and EfficientNet. Because saliency is heuristic, further causal validation using counterfactual occlusion or structure–function probing is warranted.

Uncertainty quantification further enhances safety. Tsallis entropy (q=0.25) served as a monotonic confidence measure, enabling triage of outputs by reliability. Future work could integrate Bayesian ensembling or evidential learning to better separate epistemic and aleatoric uncertainty.

Limitations include VF-based labeling noise, irregular visit spacing, class imbalance, and potential dataset shift across technicians or devices. Despite these constraints, cross-validated performance and subgroup stability indicate strong generalizability.

Future directions involve prospective, multi-center validation with clinically meaningful endpoints, time-aware sequence encoders for irregular sampling, and incorporation of additional imaging modalities (macular OCT, ONH en face, fundus photos). Domain harmonization and cost-sensitive objectives may further improve equity and real-world utility.

## V. CONCLUSION

In this large-scale retrospective cohort, multimodal sequence-aware deep learning achieved reproducibly high discrimination for forecasting glaucoma progression 2-4 years after baseline. The ConvNeXt-based model performed best (AUC ≈0.95; accuracy ≈ 0.94-0.96; F1 ≈0.85–0.88), surpassing ViT, MobileNet, and EfficientNet backbones. Performance was consistent across sex and race subgroups, with modest attenuation in participants older than 70 years. Saliency maps highlighted anatomically plausible RNFL arcuate bundles and corresponding VF arcuate/paracentral regions, while Tsallis-entropy provided a monotonic confidence measure for triaging uncertain predictions. Collectively, these results demonstrate that integrating structural OCT, functional VF maps, and clinical variables within a longitudinal modeling framework enables accurate, interpretable, and uncertainty-aware forecasting of glaucoma progression over multi-year horizons.

## Data Availability

All data produced in the present study are available upon reasonable request to the authors.

## Acknowledgment

This research was funded by the National Institutes of Health (NIH: R01 EY036222 and R21 EY035298), and MIT-MGB AI Cures Grant.

## AUTHOR Contribution

M.M.: Conceptualization, Methodology, Algorithm Design, Data Curation, Software (initial code development), Formal Analysis, Validation, Supervision, Visualization, Writing Original Draft, Review & Editing.

J.C.X.: Software (code development and model training), Visualization, Writing Original Draft (Abstract, Discussion, and Conclusion), Review & Editing.

M.E.: Validation, Visualization, Review & Editing.

M.W. and T.E.: Resources, Funding Acquisition.

N.Z.: Clinical Grading, Project Administration, Resources, Funding Acquisition, Review & Editing.

---

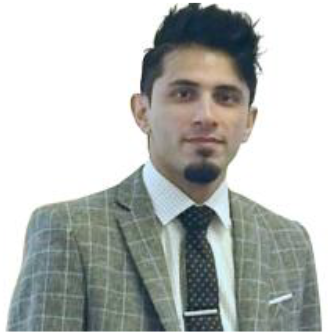

**Mousa Moradi** received the M.S. degree in biomedical engineering from Wichita State University, Wichita, KS, USA, in 2020, and the Ph.D. degree in biomedical engineering from the University of Massachusetts, Amherst, MA, USA, in 2024. Their doctoral dissertation focused on computational modeling and machine learning methods for medical image analysis and model validation in kidney assessment, AMD detection, and pulse oximetry.

He is currently a Postdoctoral Fellow with the Department of Ophthalmology, Massachusetts Eye and Ear, Harvard Medical School, Boston, MA, USA. Their research has resulted in over 30 peer-reviewed manuscripts and book chapters, including 14 first author and 2 senior-author publications. Their current research interests include AI-driven precision ophthalmology, multimodal fusion models for disease progression prediction, semi-supervised natural language processing for clinical subtyping, and the integration of imaging and genetic data for glaucoma research.

Dr. Moradi is a member of ARVO, SPIE, and Optica Open Advisory Board. They have received numerous awards including the SPIE Photonics West Best Student Paper Award Finalist, Excellence in Research Award (University of Massachusetts), Three Minute Thesis Finalist (University of Massachusetts), UMASS Dean Fellowship Award, James Southerland Garvey International Scholarship Award, and the BME MS Scholarship Award from Wichita State University.

